# LORE: A Literature Semantics Framework for Evidenced Disease-Gene Pathogenicity Prediction at Scale

**DOI:** 10.1101/2024.08.10.24311801

**Authors:** Peng-Hsuan Li, Yih-Yun Sun, Hsueh-Fen Juan, Chien-Yu Chen, Huai-Kuang Tsai, Jia-Hsin Huang

## Abstract

Effective utilization of academic literature is crucial for Machine Reading Comprehension to generate actionable scientific knowledge for wide real-world applications. Recently, Large Language Models (LLMs) have emerged as a powerful tool for distilling knowledge from scientific articles, but they struggle with the issues of reliability and verifiability. Here, we propose LORE, a novel unsupervised two-stage reading methodology with LLM that models literature as a knowledge graph of verifiable factual statements and, in turn, as semantic embeddings in Euclidean space. Applied to PubMed abstracts for large-scale understanding of disease-gene relationships, LORE captures essential information of gene pathogenicity. Furthermore, we demonstrate that modeling a latent pathogenic flow in the semantic embedding with supervision from the ClinVar database leads to a 90% mean average precision in identifying relevant genes across 2,097 diseases. Finally, we have created a disease-gene relation knowledge graph with predicted pathogenicity scores, 200 times larger than the ClinVar database.

## Main

Knowledge curation from scientific literature is fundamental for research advancement across disciplines^1–3^. Traditionally, domain-specific knowledge accumulates incrementally and relies heavily on human expert review processes. The biomedical literature, for instance, is a key resource for identifying causal genetic elements associated with diseases and offering insights into clinical practice. Several expert-curated databases, such as ClinVar^4^, COSMIC^5^, OMIM^6^, and PharmGKB^7^, provide invaluable assessments of literature evidence. Still, these resources are limited in scale due to the broad scope and the rapid expansion of scientific publications^8^. Many computational approaches have been applied to enhance automation in biomedical literature-based discovery for different tasks, such as gene-disease association prediction^9–11^, text-mining and curation^12–16^, and biomedical entity relation extraction^17–20^. Nevertheless, these methods focus primarily on extracting isolated sentences or paragraphs containing entities of interest, rather than synthesizing comprehensive information across multiple sources and contexts. Thus, substantial efforts are required to create task-specific datasets and model training for each domain of literature.

On the other hand, Machine Reading Comprehension (MRC)^21^, which requires machines to answer questions based on textual content, aims to provide a promising complement to human expertise in reading vast amounts of literature. Recent advancements in natural language processing, particularly the development of Large Language Models (LLMs), have significantly enhanced MRC capabilities to potentially accelerate knowledge synthesis^22–24^. However, while recent LLMs such as GPT-4 demonstrating remarkable capabilities in textual comprehension across diverse domains, they have difficulties in terms of reliability and verifiability^23,25^. Specifically, LLMs are prone to hallucination, a phenomenon where they generate plausible but factually incorrect information. Moreover, the opaque parametric memory of LLMs poses a substantial obstacle to traceability—the sources of evidence supporting their statements are often unclear. To work around these concerns, researchers have applied Retrieval Augmented Generation (RAG) in LLM-based chatbots^26,27^. RAG restricts the information source for LLM to an explicit but small set of retrieved texts per user query. However, due to scalability constraints, fast but shallow sentence similarity-based retrieval is required, leading to incomplete information capture as nuanced content and relevant articles are often missing.

In essence, scalable knowledge curation calls for a computational method that is inductive across domains and able to capture nuanced textual context for knowledge synthesis, all while maintaining essential reliability and verifiability. To this end, we introduce LORE (LLM-based Open Relation extraction and Embedding), a novel literature semantics framework that encompasses the best of the two worlds and is tested true in capturing disease-gene relationships across the PubMed literature (Fig. 1).

**Fig. 1.**
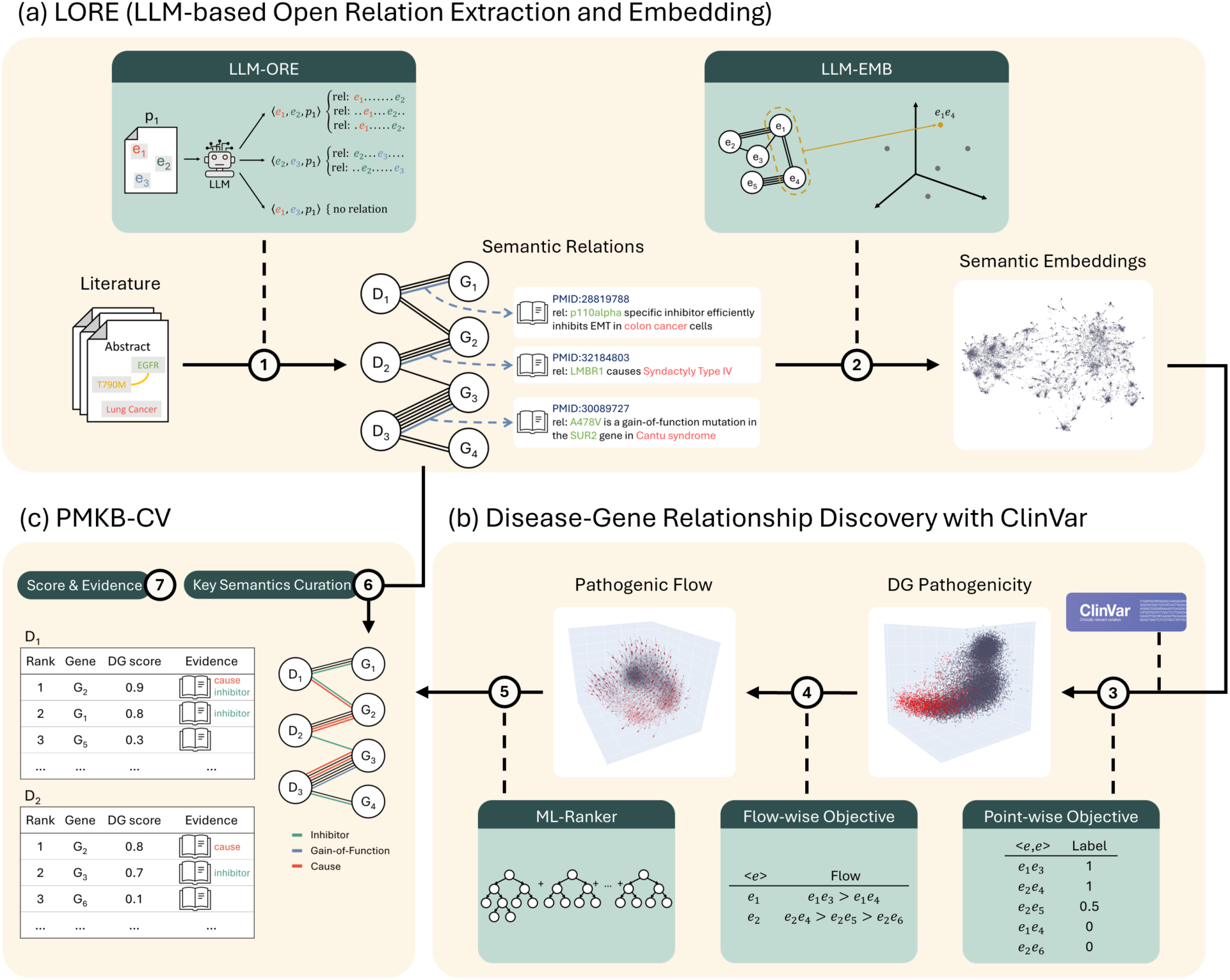
Overview of the literature semantics framework. Given a large literature containing expert domain knowledge, the framework creates a comprehensive unsupervised knowledge graph and numerical embeddings between entities. This enables large-scale supervised modeling of downstream tasks, where model predictions are accompanied by verifiable relations evidence that can be traced back to the original articles. **①** We applied the framework to the PubMed literature and created a knowledge graph containing semantic relations between diseases and genes and their mutations. **②** An embedding for disease-gene relationships is created, where each point in space contains the literature-semantic knowledge of a DG (disease-gene pair). **③** The embedding is shown to contain a latent structure of DG pathogenicity. **④** We further analyzed the disease-wise pathogenic flow and found it is cross-disease consistent and even smoother than pointwisedistribution. **⑤** An ML-Ranker is trained to model the flow and to predict pathogenic genes for each disease, where the prediction scope is 200x larger than expert-curated supervision. **⑥** We curated 105 key semantics about DG pathogenicity with linguistic lemmas to automatically tag relations. **⑦** With the proposed framework, we facilitate future research on DG pathogenicity with a literature-scale knowledge base of predicted DG scores and supporting evidence.

LORE leverages a two-stage reading methodology with the combination of LLM-based Open Relation Extraction (LLM-ORE) and LLM-based embedding (LLM-EMB) (Fig. 1a). First, LORE employs LLM-ORE to comprehend each article and generate atomic statements about entity relations therein. In other words, LLM takes charge of comprehension but the curated knowledge is explicitly derived from each article, making the generated relations reliable and verifiable.

Furthermore, this operation creates a comprehensive unsupervised knowledge graph of the literature, the conciseness of which makes it possible for LORE to then use LLM-EMB to read the full knowledge set between each entity pair and create numerical embeddings for downstream task-specific applications.

The ClinVar^4^ database is an important annotation repository of the relationships between genes and diseases. By leveraging key disease-gene pairs from ClinVar as a reference, we were able to evaluate the effectiveness of LORE to explore the latent space about gene-wise pathogenicity and disease-wise pathogenic flow across genes (Fig. 1b). In addition, we constructed machine learning models with the supervision of pathogenic genes from ClinVar to rank the relevance level of gene pathogenicity using the semantic embeddings (Fig. 1b).

Finally, we have curated a taxonomy of key semantics to use as tags for relations (Fig. 1c). We created PMKB-CV (pubmedKB-ClinVar), a novel resource to expand the scope of disease-gene relationship data. Notably, PMKB-CV encompasses more than 2,000 diseases and covers disease-gene pairs (DGs) at a scale 200 times larger than ClinVar. Moreover, PMKB-CV provides rich annotations including semantic embeddings, predicted DG scores, and verifiable knowledge graph relations with tags and source article IDs (Fig. 1c). In summary, our LORE framework harnesses the power of LLM-based MRC and enables literature-scale knowledge graph construction and downstream modeling. Importantly, PMKB-CV further bridges the gap between large-scale computational analysis and human assessment, helping to advance our understanding of disease genetics and potential therapeutic targets.

## Results

### LLM-ORE extracts semantic relations

We applied LLM-ORE to PubMed abstracts to annotate disease-gene relationships and created a comprehensive unsupervised knowledge graph (Fig. 1a). A total of 11 million relations across 1.7 million abstracts were collected by prompting GPT-3.5^28^. Using text-continuation prompts^29^, we employed LLMs to analyze individual articles and extract atomic statements describing relations between pairs of entities. Fig. 2 illustrates the prompt structure used to guide GPT-3.5 in this task. This prompt is composed of two key sections. The first section demonstrates a domain-agonistic open relation extraction (ORE). The text serves as a primer, independent of the specific biomedical context, to establish expected format and level of detail for the extraction process (see more details in Methods). Following the same structure as the demonstration, the second section applies the ORE process to the target article and entities under investigation. Notably, this approach allows for a generalized ORE from literature, not constrained to predefined relation types or entity pairs. In total, 358K distinct semantic lemmas are present across the 11M disease-gene relations.

**Fig. 2.**
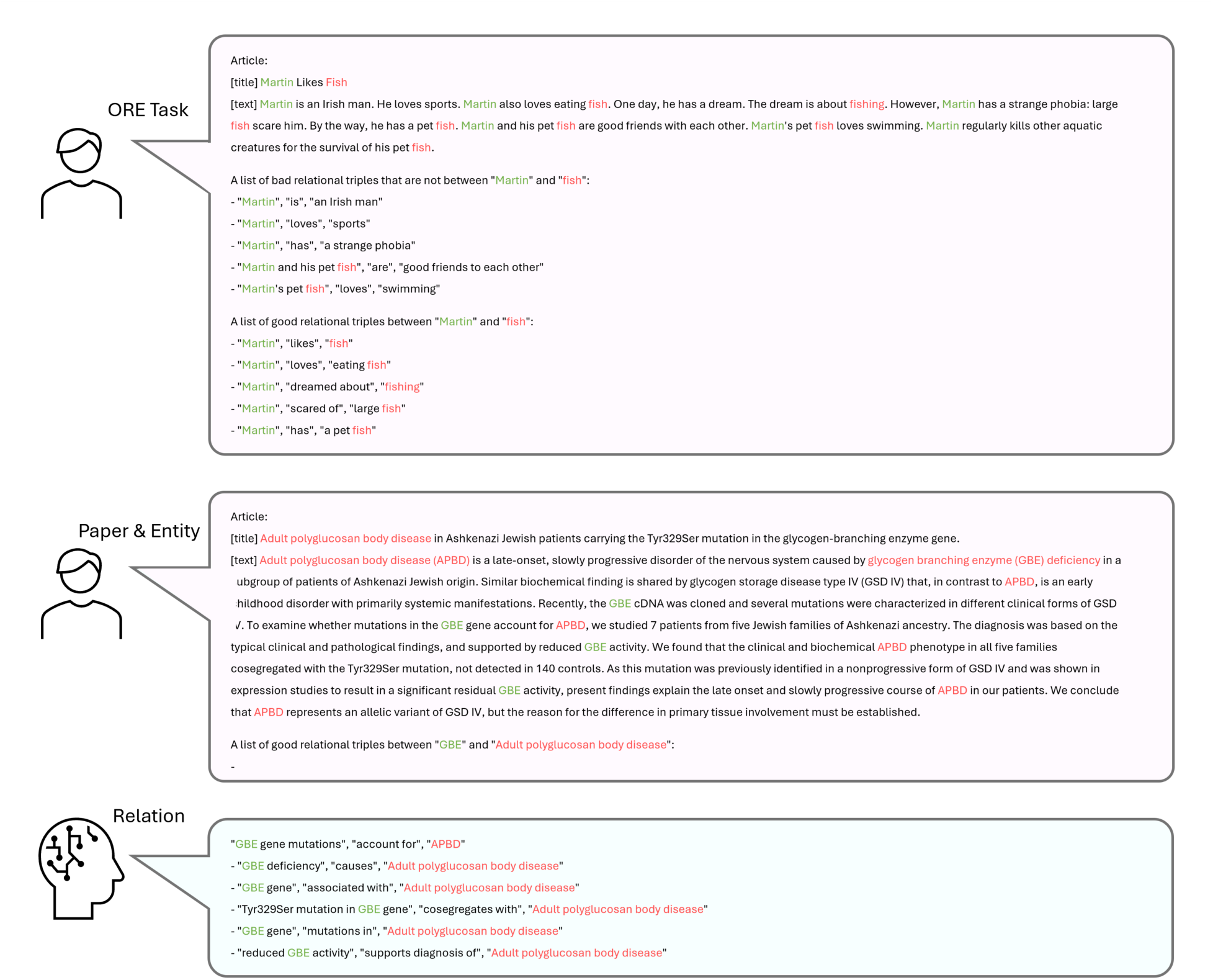
Annotating articles with LLM-ORE (open relation extraction). Text-continuation prompts are used to make LLM write down its understanding of an article as atomic statements of fact. The ORE task is crafted to extract concise relations between entities at the article level. For example, “Martin dreamed about fishing” requires comprehension and rewriting of several sentences. Also, common unwanted behaviors are avoided by providing examples of bad relations. We applied the task-agnostic prompt to extract an open set of diverse and comprehensive entity relationships for academic literature.

Next, we reviewed 282 high-coverage lemmas in the knowledge graph and curated a taxonomy of 105 key semantics about pathogenicity (Fig. 3). The key semantics are automatically tagged to relations and act as a set of indexes to access the knowledge graph. Below, we manually group key pathogenicity semantics into four main classes.

**Fig. 3.**
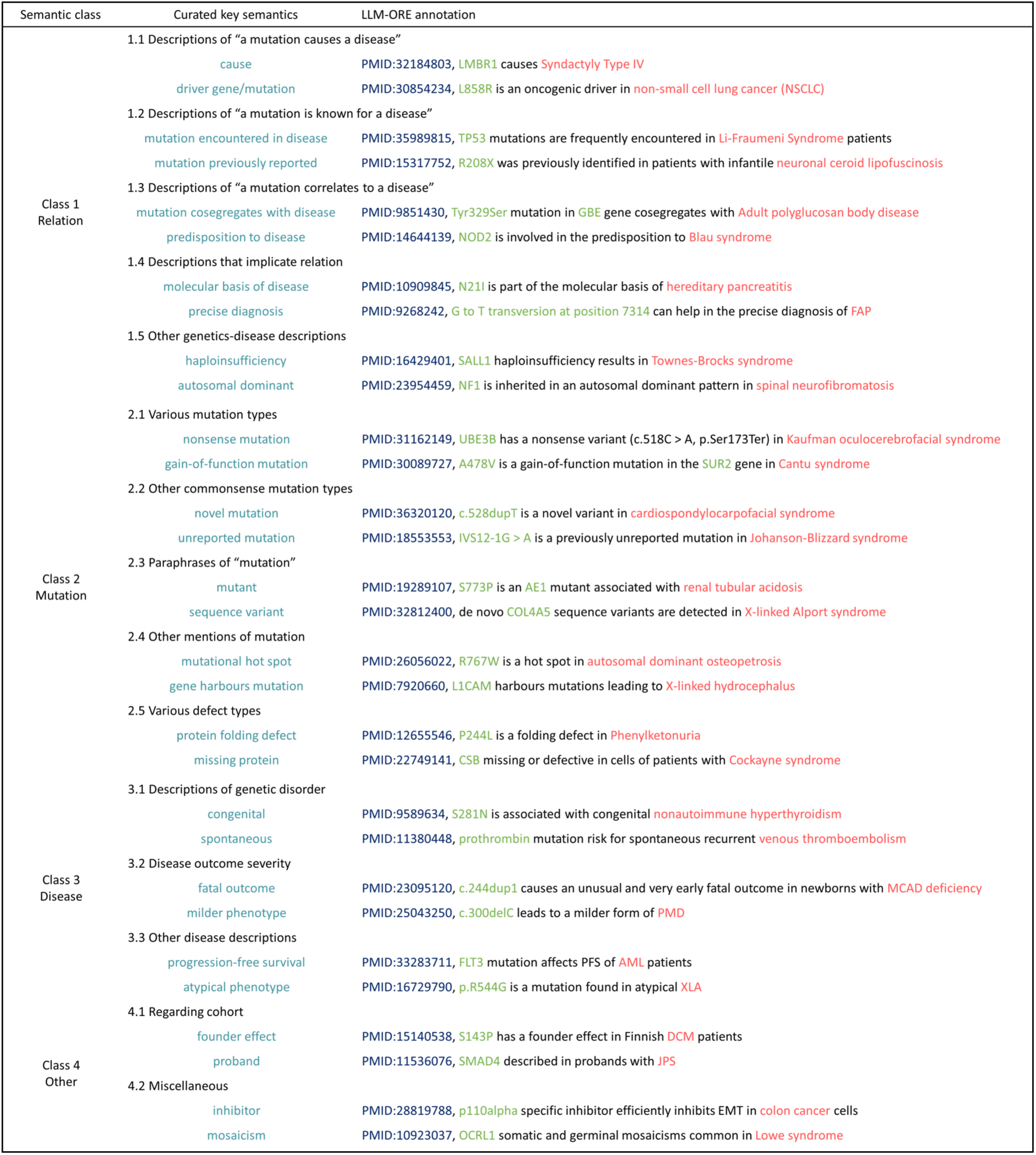
Curated key semantics. We created a taxonomy of 4 main classes, 15 sub-classes, and 105 key semantics about disease-gene pathogenicity. With their corresponding linguistic lemmas, tags are added to relations automatically. Here, two key semantics and example tagged relations are shown for every sub-class.

Class 1, Relation, includes key semantics that directly describe the relation between a gene and a disease. For example, the key semantics “mutation cosegregates with disease” in Class 1.3 describes the occurrence of a certain genetic mutation correlates to whether the genome is from a patient of a certain disease. And this is the semantics expressed by the following sentential relation.

#### LLM-ORE: Tyr329Ser mutation in GBE gene cosegregates with Adult polyglucosan body disease

Class 2, Mutation, regards the information about genetic mutations. For example, the key semantics “protein folding defect” in Class 2.5 is about a certain mutation that causes defective protein folding. This kind of information implicates that the corresponding gene has a role in a certain disease, as in the following relation.

#### LLM-ORE: P244L is a folding defect in Phenylketonuria

Class 3, Disease, on the other hand, regards genetic aspects of diseases. For example, the key semantics “milder phenotype” in Class 3.2 means a certain disease is in a milder form. This semantics implies a certain gene is at play, as seen in the following.

Class 4, Other, contains cohort and miscellaneous information that entails or hints at pathogenicity. For example, the key semantics “inhibitor” in Class 4.2 can be used to describe that a protein is a drug target, hinting at the pathogenicity of the corresponding gene:

#### LLM-ORE: p110alpha specific inhibitor efficiently inhibits EMT in colon cancer cells

### LLM-EMB embeddings capture underlying gene pathogenicity

In the second stage of LORE, we applied LLM-EMB to the PubMed DG (disease-gene pair) knowledge graph created in the first stage by LLM-ORE. For each entity pair, all their relations across all articles are encoded to a single vector, hence it contains the literature knowledge about their relationship. A dense 512-dimensional representation is created for each DG.

To analyze the latent pathogenicity structure within the literature-semantic embedding, we first display the embedding space with 2D UMAP^30^ (Fig. 4a-d). Each point represents a DG. When points are colored by PubMed abstract co-occurrence frequency, high-frequency DGs are seen distributed throughout the space (Fig. 4a). When points are instead colored by ClinVar pathogenicity labels, pathogenic DGs are seen clustering toward a subspace (Fig. 4b). This subspace is captured well by the pathogenic score prediction of ML-Ranker. With an optimal pathogenic score threshold to split the DGs and color them red and gray, the distribution is close to the ClinVar labels (Fig. 4d). Furthermore, while the ClinVar labels are human-curated and sparse, the graded pathogenic score predicted by ML-Ranker is seen to paint a smooth landscape of pathogenicity (Fig. 4c). Thus, high-score DGs not curated yet in ClinVar will be of high interest to the biomedical community.

**Fig. 4.**
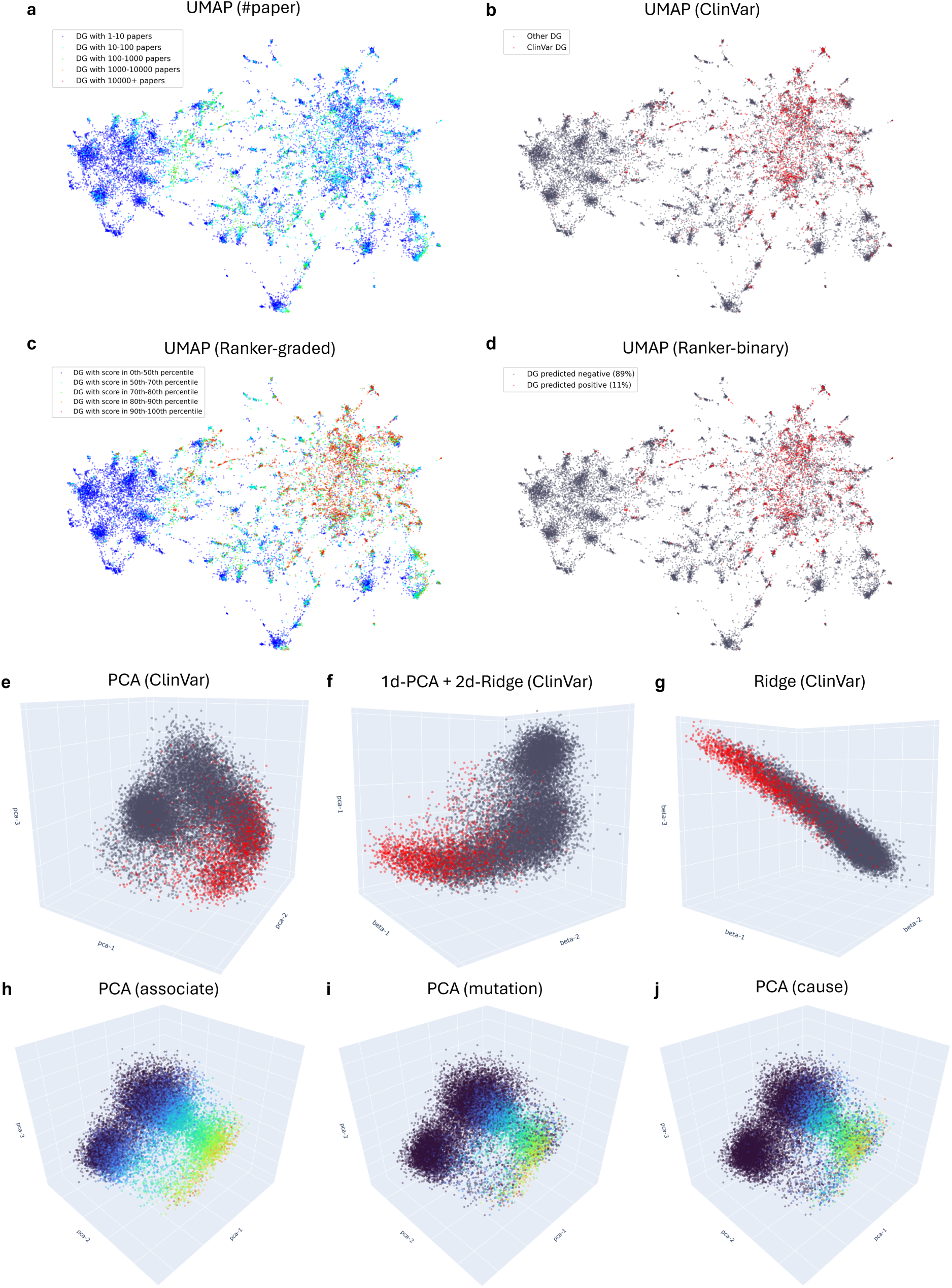
Literature-semantic embedding visualization for disease and gene associations. **a-d**, Visualization with UMAP. Points are colored by the number of papers (#paper) (**a**), ClinVar pathogenicity labels (**b**), graded ML-Ranker prediction (**c**), and binary ML-Ranker prediction (**d**). The sparse ClinVar-curated red pathogenic DGs (disease-gene pairs) are seen clustering toward a subspace and captured well by ML-Ranker, which also provides a smooth landscape of graded predictions for un-curated DGs. **e-g**, Visualization with linear axes calculated by PCA (**e**), Ridge (**g**), and their combination (**f**). A point is colored red if it is a known pathogenic DG in ClinVar, and DGs with unknown pathogenicity are colored gray. A latent structure of two manifolds—a gray ball of non-pathogenicity and a curved arm of transition from non-pathogenicity to pathogenicity—resides in the semantic space. **h-j**, Visualization with PCA and colored by distributions of literature semantics “associate” (**h**), “mutation” (**i**), and “cause” (**j**). The connection between the smooth semantics distribution and the sparsely-curated ClinVar pathogenicity distribution can be seen.

Next, we show 3D linear subspaces of the LLM-EMB embedding and observe the DG pathogenicity distribution (Fig. 4e-g). The axes are calculated by Principal Component Analysis (PCA) or ridge regression, a regularized version of Ordinary Least Squares (OLS), against ClinVar pathogenicity labels. For the PCA subspace (Fig. 4e), the top 3 dimensions explain 12.8% of the variance of the embedding. A latent structure of two manifolds can be seen—a dense gray ball of non-pathogenic DGs and a curved arm marked by a gradual transition from non-pathogenicity to pathogenicity. For the subspaces spanned by a combination of PCA axes and regression axes (Fig. 4f, g), clearer manifolds can be seen with more regression axes included. With two regression axes, a gray ball of non-pathogenicity and a gray-to-red arm of pathogenicity transition are observed. We note that the regression axes span a smaller subspace of the embedding compared to the PCA axes. Nevertheless, they provide an analytical pathogenicity perspective of the unsupervised embedding.

On the other hand, we analyzed the literature semantics structure by visualizing the distribution of important semantic lemmas in the 3D PCA subspace (Fig. 4h-j). For each semantic lemma, such as “cause”, we show its frequency in log scale per DG with a cool-blue-to-warm-red color scale. Clear patterns are observed for various lemmas, especially for lemmas that have a flatter distribution, such as “associate”, “mutation”, and “cause”. The semantics “associate” (Fig. 4h) is seen increasing along a linear axis but is more frequent (greener) in the curved arm than in the sparse parts (bluer). On the other hand, the semantics “mutation” (Fig. 4i) is distributed along the curved arm. Finally, the semantics “cause” (Fig. 4j) is also distributed along the curved arm but is much centered in the pathogenic end of the arm.

The semantic lemma visualization reveals a connection between the intrinsic semantics structure of LLM-EMB and the latent DG point-wise pathogenicity structure. The gray ball of ClinVar non-pathogenicity coincides with a lack of key literature semantics about pathogenicity. The gray-to-red arm of the ClinVar pathogenicity transition is reflected by an increase in the key literature semantics.

### Pathogenic Flow in The Literature-Semantic Space

In this section, we delve into the embedding space underlying the DG pathogenicity distribution (Fig. 5). First, to model pathogenicity, a straightforward approach would be to model the distribution of pathogenic DGs, i.e., red points, in the embedding space (Fig. 5a,b). In the literature-semantic space visualized with 3D linear axes (Fig. 4e-g), there are clear subspaces of pathogenicity and non-pathogenicity. These low-rank subspaces can be seen in the gray ball and the ends of the gray-to-red arm. But, there are also places where gray and red dots mix, such as the center of the arm. To distinguish them, high-dimensional hyperplanes or complex non-linear subspaces are needed, requiring more data and threatening generalization.

**Fig. 5.**
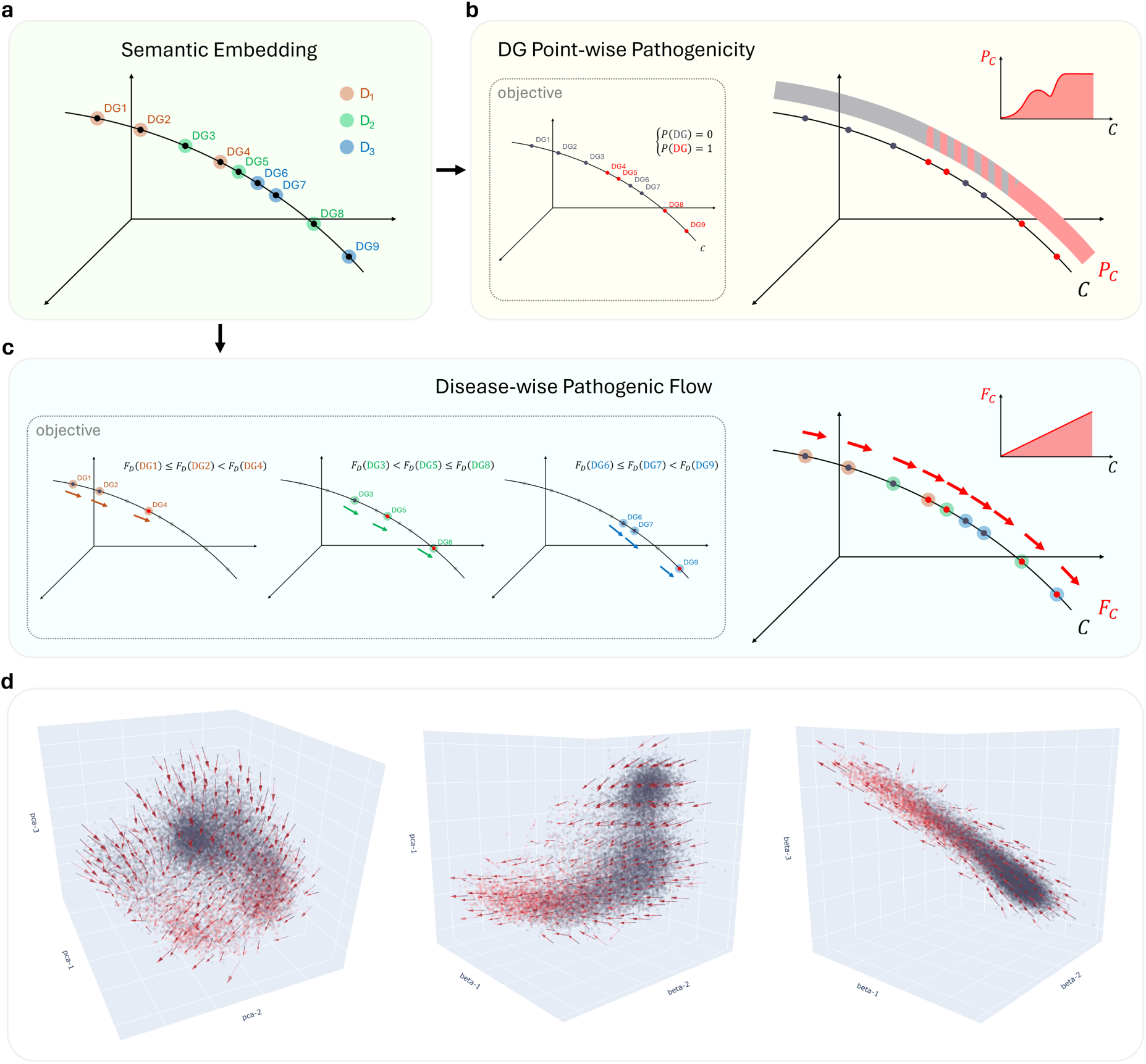
Pathogenic flow in the literature-semantic space. **a**, Suppose in the semantic embedding, 9 DGs from 3 different diseases reside on a curve. **b**, For the DG point-wise objective, the disease group information is not used, and absolute zero-one labels are the prediction target. As a result, a non-linear function along the curve must be learned. **c**, For the disease-wise flow objective, DGs are grouped by disease, and relative flow directions are the prediction target. Because of the cross-disease consistency of the flow, a linear function along the curve will be learned to rank DGs for every disease perfectly. **d**, Visualization of the actual pathogenic flow. A smooth, cross-disease consistent field of pathogenic flow is seen residing in the literature-semantic space.

However, the fundamental task is to predict the most relevant pathogenic genes for each disease. For each disease, if its gray and red points are distributed separately to the two ends of a linear axis or, more generally, a smooth curve, pathogenic and non-pathogenic genes can be distinguished by splitting the curve. Furthermore, if the curves are consistent across diseases, the relevant pathogenic genes for every disease can be identified by monotonically raising predicted relevance along the curves. Under this condition, perfect modeling can be achieved even if non-pathogenic and pathogenic DGs from different diseases are mixed in the embedding space (Fig. 5c).

In this work, we define pathogenic flow as the direction and magnitude of non-pathogenicity to pathogenicity at each location in space. We start by calculating the disease-wise flow direction at each DG. Then, we quantize the flow direction at each location in the space, using flow magnitude to reflect consistency. We observed a smooth, cross-disease consistent field of pathogenic flow in the literature-semantic embedding (Fig. 5d). Along the arm-shaped manifold of gray-to-red transition, there is a consistent flow of non-pathogenicity to pathogenicity. This means that despite DG point-wise modeling being difficult in the center mixture of the arm, disease-wise modeling is much more linear and smoother.

### Literature-Scale Pathogenicity Prediction by ML-Ranker

Here, we focus on building an ML-Ranker that models disease-wise pathogenic flow and can score every DG that co-occurred in PubMed abstracts. We created PMKB-CV (Fig. 6a), a dataset containing 2,097 diseases that are in both pubmedKB^2^ and ClinVar^4^.

**Fig. 6.**
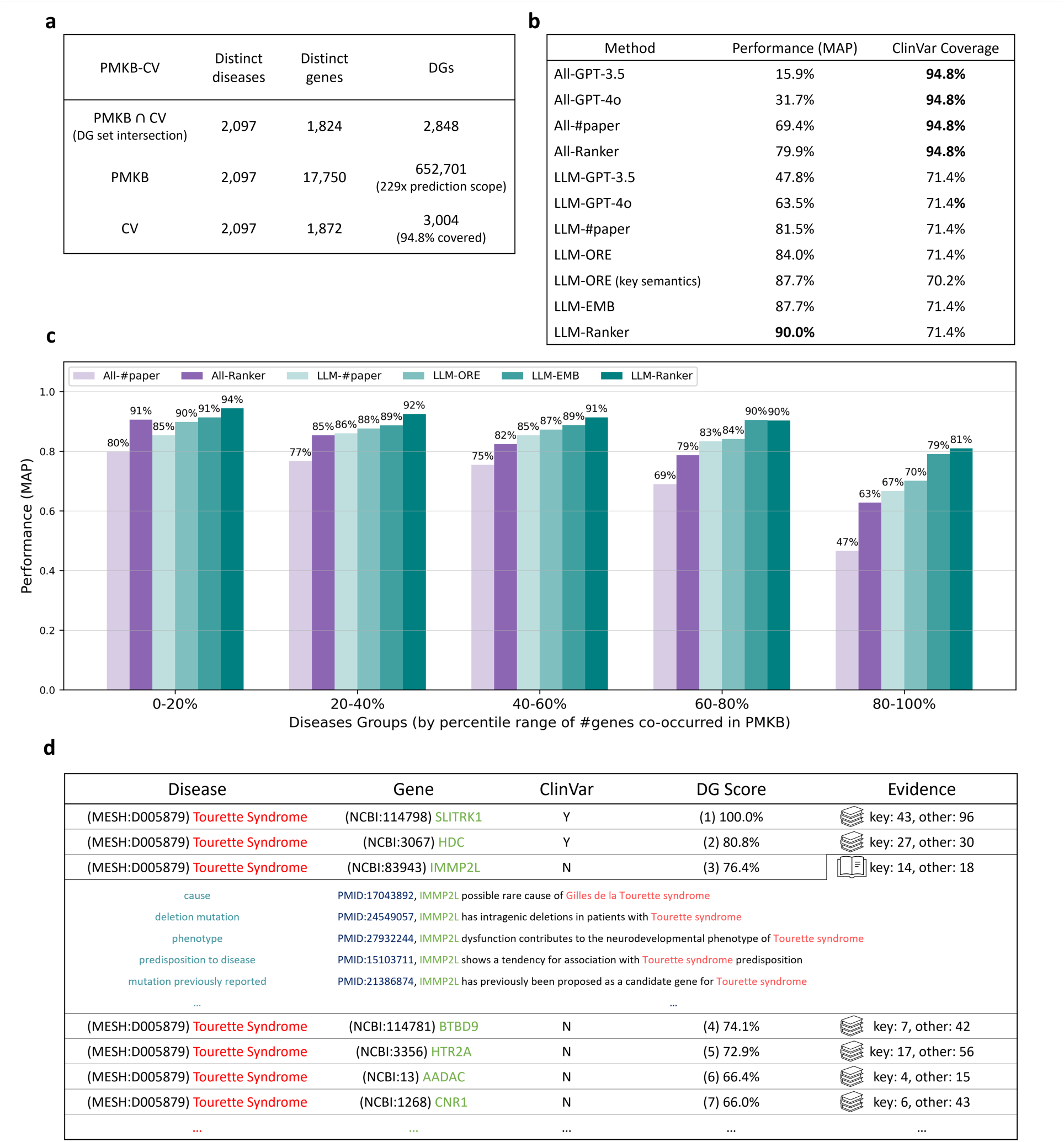
The PMKB-CV dataset and ranking performance. **a**, The statistics of the PMKB-CV dataset. For 2K diseases, the literature semantics framework has a 200x prediction scope against curated DGs. **b**, The ranking performance MAP (mean average precision) and the ClinVar coverage of different methods. GPT-4o does not perform well. In comparison, LLM-EMB linear regression alone achieves 87.7% performance, while the full-fledged ML-Ranker provides higher performance at 90.0% or higher coverage at 94.8%. **c**, Performance across diseases of different scopes. While naive paper counting has difficulty ranking DGs for those diseases that co-occurred with many genes in PMKB, our semantic embedding approach remains robust. **d**, Top-ranked DGs, 7 out of 586 in PMKB, for Tourette syndrome accompanied by their literature relation evidence.

We applied the lambda objective with Gradient-Boosted Decision Trees (GBDT)^31^ to directly model disease-wise pathogenic flow. Ordinary DG point-wise regression optimizes the absolute pathogenicity score (Fig. 5b). Instead, the lambda objective optimizes disease-wise ranking by contrasting pathogenic and non-pathogenic genes on a per-disease basis (Fig. 5c). The relaxed objective is plausible because of the smoothness and cross-disease consistency of the latent pathogenic flow in the literature-semantic space (Fig. 5d).

We compiled the PMKB-CV dataset to validate the proposed approach (Fig. 6a). PMKB-CV contains 2K diseases that are in both pubmedKB and ClinVar. For these diseases, 600K DGs co-occur in PubMed abstracts while only 3K DGs are human-curated by ClinVar. For the 3K known pathogenic DGs, paper abstract co-occurrence covers 94.8% and LLM-ORE relations cover 71.4% (Fig. 6b). We include pubmedKB annotations, such as the number of paper abstracts in which a DG co-occurs, in the dataset and also use them as model features. And, we used zero embedding for those DGs without relations. As a result, all DGs in the dataset can be uniformly used for training and prediction.

By using leave-one-disease-out cross-validation, we could evaluate the performance of ML-Ranker on prediction of pathogenic genes for each disease iteratively. An average precision (AP) score is calculated for each disease, and mean average precision (MAP) is used to evaluate the overall performance of the ranker. As a baseline, we directly asked OpenAI GPT-3.5 and the latest GPT-4o about the pathogenicity of each DG. In addition, to put the effectiveness of the curated key semantics in identifying crucial literature evidence into perspective, we tested a DG pathogenicity prediction method of simply counting the number of tagged relations per DG.

For all DGs in PMKB-CV, ML-Ranker achieves 79.9% MAP, a leap from the 69.4% MAP of paper counting and the 31.7% MAP of GPT-4o. If we narrow the scope to specifically those DGs with LLM-ORE annotations, 90.0% MAP is achieved by the full-fledged ML-Ranker. Performance steadily improves across paper counting, literature-semantic relation counting, linear regression on literature-semantic embedding, and latent pathogenic flow modeling (Fig. 6b). Furthermore, for the most popular diseases that co-occurred with the most genes in PMKB, ML-Ranker achieves a robust 81% MAP performance compared to the 67% MAP of paper counting (Fig. 6c). Finally, the DG scores and ranking provided by ML-Ranker are accompanied by literature evidence (Fig. 6d). We facilitate future expert assessment of DG pathogenicity by a quick grasp of literature knowledge with key semantics relations and relevant articles.

## Discussion

Recent advancements in LLMs aim to automate complex sensemaking as human endeavors in reading and connecting information across large collections of scientific literature^32^. Our study introduces LORE, a novel literature semantics framework that fundamentally reframes how we leverage LLMs to extract and utilize knowledge from scientific literature. The framework offers several key advantages.

First, knowledge synthesis using the LORE approach constructs a literature knowledge graph of verifiable factual statements linked to the sources. Second, LORE offers a scalable framework for knowledge synthesis from large amounts of article texts. LORE extracts original article texts and transforms them into a concise knowledge graph. This is more efficient than using a traditional RAG^26^ approach, where only a small set of articles is selected for LLM to read. The knowledge graph is much more concise compared to the original articles. This reduction in size and complexity allows for a more efficient representation of information, thereby all the relevant knowledge can then be embedded for downstream tasks. In addition, LORE allows new publications to be annotated and expands the knowledge graph continually. Third, this approach places much less demand on the capability of LLMs compared to directly asking LLMs expert domain questions. Using LORE, we have captured gene pathogenicity with GPT-3.5, a feat far from being achieved by directly asking GPT-4o (Fig. 6b). Indeed, as much smaller and open-source LLMs have been shown competent for article-level comprehension^33,34^, the methodology is not constrained to enterprise LLMs. Finally, while our framework demonstrates remarkable efficacy in capturing disease-gene relationships through unsupervised relationship extraction and embedding, users could also employ LORE with prompt engineering and fine-tuning to annotate task-specific knowledge across various domains of scientific inquiry^35,36^.

When applying LORE to the complex landscape of DG relationships, we unveiled that a latent smooth field of cross-disease consistent pathogenic flow is present in the unsupervised literature-semantic embeddings. This discovery reveals that although pathogenic and non-pathogenic DGs from different diseases may occupy similar locations in the embedding space, there exists a consistent directional flow of pathogenicity in terms of semantics. To illustrate, consider a simplified one-dimensional embedding axis where rare and common diseases coexist (Supplementary Fig. 1). Suppose D1 is a rare disease and is reported in few studies, while D2 is a common disease, and a multitude of papers discuss its possible association with many genes. Then the following literature annotations, DG locations on the embedding axis, and pathogenicity labels are possible.

G1 is not related to D1. (x=0, y=non-pathogenic)
G2 mutation is found in a D1 patient. (x=1, y=pathogenic)
G3 mutation is found in a D2 patient; G3 is associated with D2. (x=2, y=non-pathogenic)
G4 mutation is found in a D2 patient; G4 causes D2. (x=3, y=pathogenic)

While the pathogenic D1G2 and the non-pathogenic D2G3 are mixed in the middle of the axis, literature evidence about pathogenicity consistently increases along the axis. As a result, even when pathogenic and non-pathogenic associations are interspersed due to inter-disease differences such as popularity, the literature-semantic axis provides for a cross-disease consistent linear flow, enabling accurate pathogenicity modeling across diseases.

Our initial analyses of this pathogenic flow revealed clusters of disease-specific pathogenic curves. Intriguingly, we found that these clusters often form a continuum, with the endpoint of one cluster serving as the starting point for another. This observation suggests a broader, interconnected field of pathogenic relationships across diseases, offering new perspectives on the complex landscape of genetic pathogenicity.

For the study of DG pathogenicity, we compiled the PMKB-CV dataset, which contains 2K diseases that are in both pubmedKB^2^ and ClinVar^4^. While only 3K DGs are human-curated in ClinVar for these diseases, our PMKB-CV dataset collects 600K DGs co-occurred in PubMed abstracts. This dataset has a vast potential for literature-based discovery. We note that the full pubmedKB contains 8K diseases and 3M DGs with article abstract co-occurrence, while the full ClinVar contains 3K diseases and 4K curated DGs. With full pubmedKB, the potential prediction scope is even larger, but there is still an inherent limit of co-occurrence to article abstract-based MRC.

In conclusion, our works make three significant contributions to the field. First, we present a novel literature semantics framework that addresses the long-standing challenges of comprehensiveness, reliability, and verifiability in MRC. Second, we demonstrate the efficacy of LORE in capturing complex pathogenic relationships across diseases to reveal new insights into the pathogenic flow. Finally, we provide a literature-scale dataset that not only complements existing resources like ClinVar but also offers a knowledge graph of DG relationships with graded pathogenicity scores for genetic prioritization in clinical practices. The methodology of LORE is a general improvement to LLM-based MRC and paves the way for bridging vast literature resources and actionable scientific knowledge for accelerated discoveries across scientific disciplines.

## Methods

### Two-Stage Reading Comprehension of LORE

In the first stage of LORE, we applied LLM-ORE by prompting LLMs. Fig. 2 shows the actual prompt we used. The demonstration consists of an article with a topic as Martin Likes Fish, target entity pair “Martin” and “fish”, along with lists of unwanted and desired annotations. This section implicitly specifies the ORE task and the following required properties.

1. *The annotations should be concrete statements of fact implied by the article*.
2. *The annotations should follow a structured format, specifically a list of relational triplets*.
3. *Each relational triplet should be <“subject”, “predicate”, “object“>*.
4. *The subject should contain the first entity, and the first entity should only appear in the subject*.
5. *The object should contain the second entity, and the second entity should only appear in the object*.
6. *The predicate should be a concise, self-contained description of the relation between the subject and the object*.
7. *Abstract understanding of the article is allowed beyond sentence-level syntax*.

The approach allows for abstract understanding beyond sentence-level syntax. The requirements are better understood through demonstration rather than explicit definitions. For instance, instead of explaining the terms “subject,” “predicate,” and “object,” the examples and counterexamples in the demonstration make the concept clear. In addition, the examples illustrate behaviors that are easier to grasp instinctively rather than by complex rules. Examples include the removal of “also” from “Martin also loves eating fish”, the rewriting of “large fish scare him” to “scare of”, and the digested understanding of “Martin dreamed about fishing” from “he has a dream. The dream is about fishing”. Finally, we note that the ending “-” is important in ensuring that LLM follows the desired list format.

Formally, the desired generation

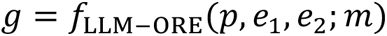

where *p* is the target article, *e*_1_ and *e*_2_ are the names of the target entities, and *m* is the LLM model. In this work, we use the paper title and abstract as *p*. For *m*, gpt-3.5-turbo-0613 is used. The function maps < *p*, *e*_1_, *e*_2_ > to *g*, the list of parsed relational triplets, using the text-continuation prompt shown in Fig. 2.

In the second stage of LORE, LLM reads all relations between an entity pair and produces a numerical representation of knowledge about their relationships. For example, the following relations between the entity *e*_1_ and the entity *e*_2_ are read by LLM as the following document.

*“e1”, “causes”, “e2“*

*“e1 mutations”, “are frequently encountered in”, “e2 patients“*

*“e1 haploinsufficiency”, “results in”, “e2“*

*Document: “e1 causes e2. e1 mutations are frequently encountered in e2 patients. e1 haploinsufficiency results in e2.“*

If the document is larger than the allowed context of an LLM, it is split into multiple sub-documents, each of which contains as many relations as possible.

Formally, the embedding vector of an entity pair is given by

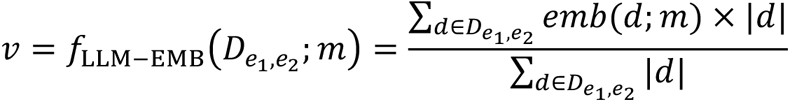

where *e*_1_ and *e*_2_ are the target entities, 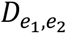 is the set of all sub-documents, usually just one full document, containing the relations between *e*_1_and *e*_2_, and *m* is the LLM model. In this work, we employed the text-embedding-3-large from OpenAI for *m*, and the length of each sub-document |*d*| is its number of tokens according to *m*.

### Modeling the Pathogenic Flow with ML-Ranker

To visualize the disease-wise pathogenic flow, we define the flow as a unit vector at each DG that points to the pathogenic direction of disease D at the embedding location of DG. Formally, the flow vector is given by

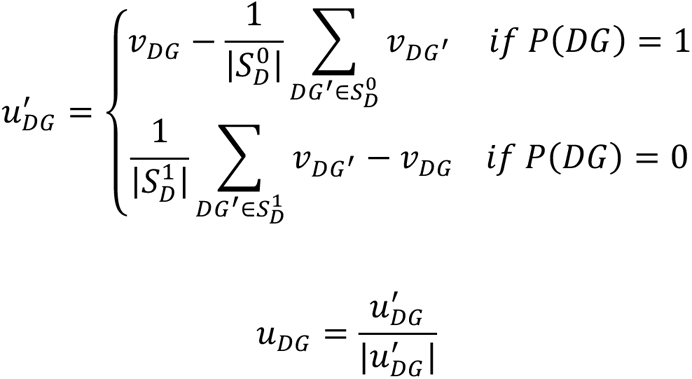

where *v* denotes the embedding vector by LLM-EMB, 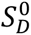 and 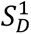 denote the sets of non-pathogenic and pathogenic disease-gene pairs of *D* respectively, and *P* maps non-pathogenic and pathogenic pairs to 0 and 1 respectively. In other words, each non-pathogenic DG has a unit flow vector directed at the average embedding of the pathogenic DGs of the same disease, and each pathogenic DG has a unit flow vector directed from the average embedding of the non-pathogenic DGs of the same disease. Then, we quantize the flow vectors for each cube in space by averaging them. Formally, a quantized flow vector is given by

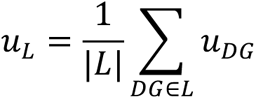

where *L* is the set of DGs in a cube subspace. Finally, we show the field of pathogenic flow in Fig. 5d, where each arrow corresponds to a quantized flow vector. The arrow direction is the aggregated disease-wise flow direction from non-pathogenicity to pathogenicity, and the arrow length, proportional to |*u_L_*|, reflects the degree of cross-disease consistency at that location.

As shown in Fig. 5c, the essence of modeling the pathogenic flow is to model the difference between non-pathogenic and pathogenic genes per disease. Specifically, suppose *DG*1 and *DG*2 correspond to the same disease but *P*(*DG*1) = 1 and *P*(*DG*2) = 0. Let *s* denote the predicted pathogenicity score. Then one would want to maximize the probability Pr (*P*(*DG*1) > *P*(*DG*2)) by minimizing the following RankNet^37^ loss.

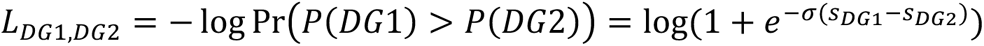

To make sure the most relevant genes are ranked on top for each disease, the final gradient, *λ*, is the gradient of the RankNet loss multiplied by the change in Normalized Discounted Cumulative Gain (NDCG)^38^. Formally, this LambdaRank^39^ gradient is given by

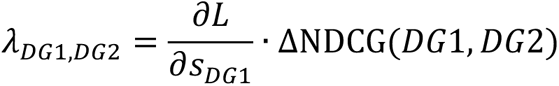

In this work, we use *λ*GBDT, the lambda objective combined with Gradient-Boosted Decision Trees (GBDT)^31^, as the ML-Ranker to explicitly model the pathogenic flow in the literature-semantic space.

To evaluate the ranking performance for each disease, average precision (AP) is used to see if the known pathogenic genes for a disease are ranked on top. Formally, for each disease,

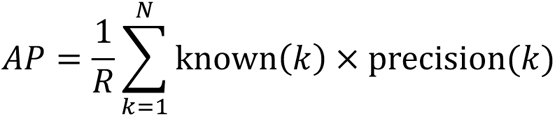

where *N* is the number of ranked genes for that disease, and *R* is the number of ranked known pathogenic genes for that disease. known(*k*) = 1 if the gene ranked at *k* is a known pathogenic gene for that disease; otherwise known(*k*) = 0 . precision(*k*) is the percentage of known pathogenic genes among genes ranked top-*k* for that disease.

### Key Semantics Curation

The curation process consists of three main steps: linguistic lemma extraction, important lemma identification, and manual taxonomy construction. In the first step, the sentential relations extracted by LLM-ORE are tokenized to bags of words, and word inflections are lemmatized to dictionary form. For example, *cause*, *causes*, *caused*, and *causing* all correspond to the same linguistic lemma. At this stage, entity names are also filtered out.

In the second step, important lemmas are identified using a coverage filter and a precision filter. The coverage filter demands a lemma to appear in LLM-ORE relations of at least *n* DGs. The precision filter requires that a certain proportion *r* of all the relations involving a lemma be from known pathogenic DGs, as indicated by ClinVar. The parameters can be adjusted according to the desired scope of key semantics. In this work, we selected a coverage parameter *n* = 100 and a precision parameter *r* = 50% and resulted in 282 important lemmas.

The final step involves manually curating a taxonomy of 105 key semantics by examining the LLM-ORE relations associated with these important lemmas During curation, we sampled 10 relations from known pathogenic DGs and 10 relations from other DGs to inspect for each lemma. The resulting key semantics were then used to tag all relevant relations in the respective lemmas. As a result, the LLM-ORE knowledge graph contains sentential relations linked to the semantic taxonomy of pathogenicity.

Furthermore, we experimented with a DG pathogenicity prediction method where the number of tagged relations for each DG is directly used as its pathogenic score. We note that the curation process has used the ClinVar information, so the generalizability of the ranking performance of this method is not directly comparable to other methods. Nevertheless, we put the effectiveness of the curation method into perspective, showing what performance and scope DG pathogenicity researchers can expect when using the key semantics tags to grasp the literature knowledge and identify relevant relations and articles for their DGs of interest.

### Constructing the PMKB-CV Dataset

We constructed the PMKB-CV dataset as a large-scale complement of ClinVar and an evaluation from Medical Subject Headings (MeSH), a vocabulary thesaurus maintained by the Nation Library of Medicine (NLM), and *Homo sapiens* protein-coding gene IDs from the National Center for Biotechnology Information (NCBI).

The PMKB-CV dataset comprises two main components including literature-based and expert database parts. For the literature part, we utilized the annotations from pubmedKB^2^. The gene and variant mentions are both indexed by NCBI gene IDs, and we considered the occurrence of either a gene or its variant as an occurrence. This approach yielded 3,128,402 DG pairs with co-occurrence within abstracts, encompassing 8,894 diseases and 18,393 genes. In addition, we extracted a subset of PubMed articles as the most relevant literature for the study of disease-gene relationships via a bootstrapping iteration with pubmedKB annotations as features. Using leave-one-disease-out training, we predicted a bootstrap score for each DG. Then, the literature subset is formed by the articles associated with the top three genes for each disease, the top three diseases for each gene, and the top 15K DGs. The resulting subset contains 1,745,538 articles, for which we applied LORE and curated 11,285,095 relations.

The expert database part was derived from ClinVar^4^, which provides gene-disease relationship data. As ClinVar uses OMIM (Online Mendelian Inheritance in Man) numbers for disease indexing, we applied UMLS (Unified Medical Language System)^40,41^, a vocabulary alignment dataset maintained by NLM, to map OMIM numbers to MeSH IDs. This process resulted in 4,311 known pathogenic DGs, spanning 3,175 distinct diseases and 2,416 distinct genes.

The final PMKB-CV dataset was created by including those diseases that have both pubmedKB co-occurrence DGs and ClinVar known pathogenic DGs. Statistics of the resulting dataset are shown in Fig. 6a.

## Data Availability

The PMKB-CV datasets supporting the findings of this study are available at https://drive.google.com/file/d/1rGgZmUOU0XIQtV3mtYsMU-4t2lJQNOfo.

## Code Availability

The code supporting the conclusions of this study is available on GitHub at https://github.com/ailabstw/LORE.

## Author contributions

P.H.L. conceived this study. P.H.L. and Y.Y.S. implemented the pathogenic flow modeling and created figures. P.H.L. and J.H.H. wrote the manuscript. H.K.T., C.Y.C., and H.F.J. helped conceptualization and in the preparation of manuscript. J.H.H. contributed to conceptualization, writing, and project supervision. All authors are involved in discussion and finalization of the manuscript.

