## Supplemental Figure 1 for "LORE: A Literature Semantics Framework for Evidenced Disease-Gene Pathogenicity Prediction at Scale"

**Supplementary Figure**

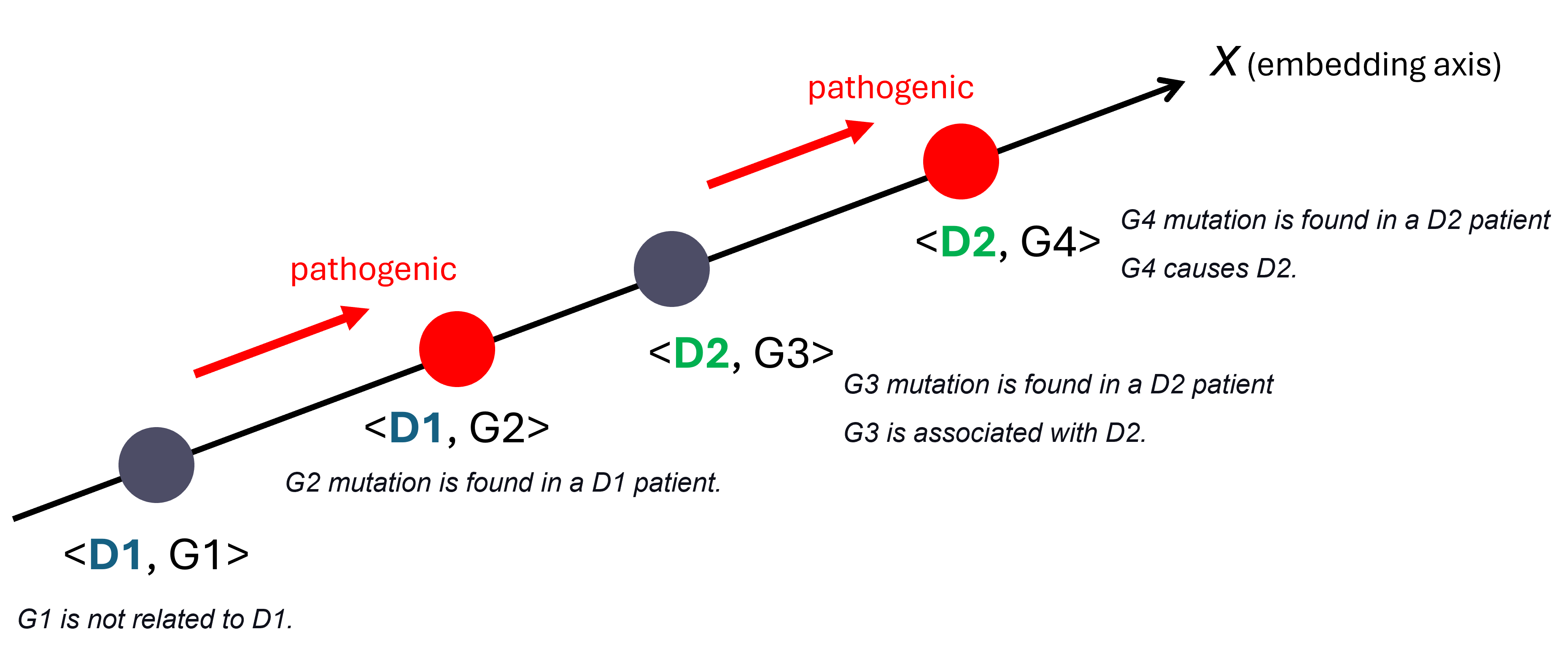
 **Supplementary Fig. 1 | Schematic pathogenic flow in the literature-semantic axis.** The axis embeds a linear pathogenic flow that can be modeled to report the most relevant genes for both diseases.
